# Multi-omic profiling of atherosclerosis: protocol and pilot data for the AtherOMICS biobank

**DOI:** 10.1101/2025.06.17.25329773

**Authors:** Luka Živković, Roya Batool, Julian Louma, Paulo Vinicius Gil Alabarse, Yingle Li, Lanyue Zhang, Stefan Mayrhofer, Anushree Ray, Mohamad Ali Antabi, Panagiotis Zangas, Jana Mattar, Anna Kopczak, Andreas Schindler, Paul Reidler, Yaw Asare, Steffen Tiedt, Lars Kellert, Barbara Rantner, Martin Dichgans, Nikolaos Tsilimparis, Marios K. Georgakis

**Author notes:** **Correspondence to:** Marios K. Georgakis, MD, PhD.

## Abstract

Omics technologies enable deep profiling of human atherosclerosis, but have mostly been applied in small studies lacking integration with other data modalities. AtherOMICS is a biobanking project linking multi-omic atherosclerotic plaque phenotyping with blood biobanking, in vivo imaging, and clinical data. Since August 2022, 246 patients scheduled to undergo carotid or femoral endarterectomy at LMU Klinikum (Munich, Germany), have been enrolled (median age 73 years, 32% female). Plaques undergo systematic histological characterization of lipid core, calcification, intraplaque hemorrhage, fibrous cap, macrophages, and smooth muscle cells. Single-nuclei RNAseq (n=17) has highlighted macrophages as the dominant intraplaque cell type, whereas paired plaque and plasma affinity-based proteomics (n=88) quantified 2,841 shared proteins showing low plaque-plasma correlation (median ρ=0.10). Cross-modality image integration (histology, *ex vivo* MRI, *in vivo* MRI, CTA) proved feasible. Taken together, AtherOMICS enables multimodal characterization of human atherosclerosis aiming for the discovery of athero-specific drug targets, molecular signatures of atheroprogression, and non-invasive biomarkers of plaque vulnerability.

## Introduction

Atherosclerotic cardiovascular disease remains the leading cause of mortality and adult disability worldwide.^1, 2^ Despite the widespread adoption of prevention strategies targeting systemic vascular risk factors, the incidence of atherosclerotic cardiovascular disease is rising. While 25-30% of ischemic strokes are directly attributable to atherosclerosis, its prevalence is high in all stroke patients and plaque burden is associated with adverse cardiovascular events and mortality.^3, 4^ Studies utilizing granular etiological subtyping and improved vascular imaging protocols have suggested that 31% of cryptogenic strokes and 49% of transitory ischemic attacks could be attributable to atherosclerosis.^5, 6^ In addition, atherosclerotic strokes are associated with increased risk of recurrence from the acute phase up to five years after the index event.^7, 8^

Decades of research have identified cellular and molecular hallmarks of atheroprogression and plaque destabilization,^9, 10^ which could reveal novel therapeutic targets, as evidenced by the development of anti-inflammatory atheroprotective strategies for the prevention of ischemic stroke and myocardial infarction.^11–13^ However, the mechanisms driving atheroprogression in humans remain underexplored.^14, 15^ Most mechanistic studies leverage animal models of atherosclerosis, which often fail to translate to humans.^16^ ^17, 18^ Current concepts of human atheroprogression are largely informed by histopathological studies conducted in the 1960s and 70s^19, 20^. Since then, significant advances in biotechnology have uncovered previously unknown molecular regulators of atheroprogression.^21–24^ Deep, multi-omic phenotyping could offer an avenue for dissecting these mechanisms and informing future trials.^25, 26^

Advancing therapies that target plaque biology requires diagnostic and risk stratification tools capable of capturing local plaque progression.^27^ While best practices emphasize the need for personalized risk assessment, existing risk estimators build on systemic cardiovascular risk factors rather than plaque progression biomarkers.^28^ Ultrasonography, CT angiography (CTA) and magnetic resonance imaging (MRI) can reliably identify atherosclerotic plaques and assess basic composition features.^29^ Similarly, omics technologies could help detect circulating signatures of plaque destabilization ^24^ for targeted therapies and treatment response monitoring.^30^ However, efforts to link *in vivo* imaging and circulating molecular signatures with extensive phenotyping of the plaque microenvironment remain limited.^31^

To address these gaps, we have launched AtherOMICS, a biobanking study that integrates deep phenotyping of human atherosclerotic plaques with peripheral blood biobanking, multimodal *in vivo* non-invasive imaging, and clinical data collection. We perform extensive histopathological and omics analyses in plaque samples from patients undergoing carotid or femoral endarterectomy and integrate the derived data with matched preoperative blood samples and *in vivo* plaque imaging. The objectives of AtherOMICS are: To (1) identify druggable molecular mechanisms driving plaque progression; (2) detect molecular signatures of plaque destabilization; and (3) develop circulating and imaging biomarkers of plaque vulnerability.

## Methods

### Study overview

A detailed version of the study protocol and procedures of the AtherOMICS study is available in the **Supplementary Methods**. The reporting of pilot data follows the STROBE guidelines for cross-sectional studies.^32^ AtherOMICS has been approved by the Ethics Committee of the Faculty of Medicine of the Ludwig-Maximilians-Universität (LMU) in Munich, Germany (approval numbers: 121-09, 22-0135, 23-0772). All participants have provided written informed consent and the study conforms to the 2008 Declaration of Helsinki.

Recruitment for AtherOMICS began in August 2022 and includes (**Fig. 1**):

- Plaque biobanking: Plaque material removed during carotid or femoral endarterectomy is collected and processed for histology, *ex vivo* MRI, and omics analyses.
- Blood biobanking: Peripheral blood is collected from eligible participants before undergoing endarterectomy.
- Clinical data collection: Demographic, anthropometric, medical and medication history, and laboratory data are collected from eligible participants and their medical records.
- *In vivo* imaging: CTAs and brain MRIs are obtained from clinical routine diagnostics and high-resolution carotid MRI scan prior to surgery.

**Figure 1.**
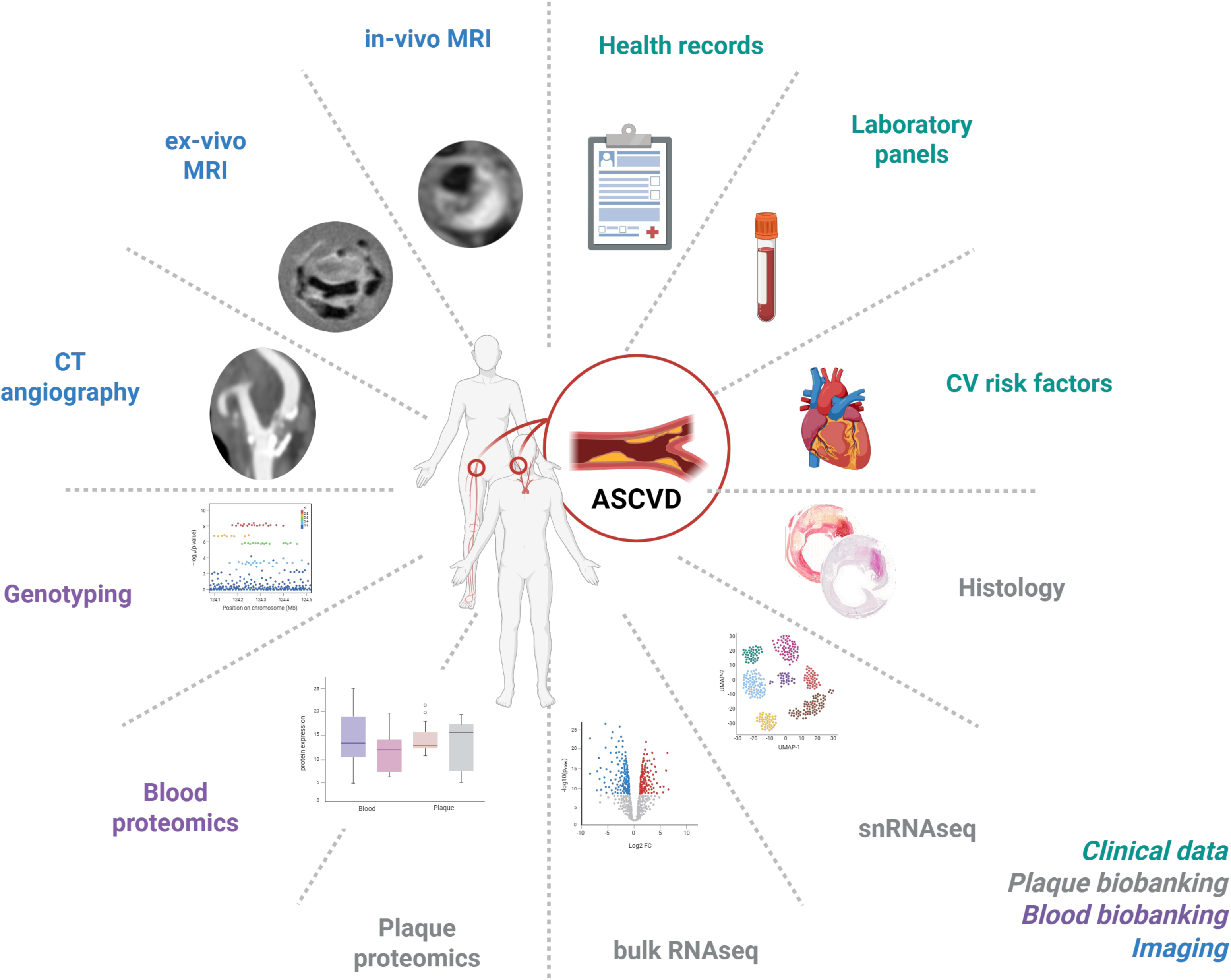
Schematic overview of AtherOMICS data types. CV = cardiovascular, snRNAseq = single-nuclei RNA sequencing, CT = computed tomography, MRI = magnetic resonance imaging. Created with biorender.com

### Study participants

The target population of AtherOMICS includes adults scheduled to undergo carotid or femoral endarterectomy due to atherosclerotic disease. Participants are consecutively recruited from the Vascular Surgery and Neurology Departments at LMU Klinikum in Munich, Germany. Reasons for referral are symptomatic or asymptomatic carotid artery stenosis and symptomatic peripheral artery disease due to stenotic lesions in the femoral artery. Study team members identify potentially eligible participants by daily screening of hospital admission records and surgery plans. The inclusion criteria for the biobank are:

1. Adults (age ≥ 18 years)
2. Scheduled for carotid or femoral endarterectomy in the next 7 days
3. Provision of written informed consent for the study prior to participation.

The exclusion criteria are:

1. Inability or unwillingness to provide written informed consent
2. Prior ipsilateral endarterectomy or stenting
3. Prior local radiation therapy.

Consenting participants scheduled for carotid endarterectomy are additionally invited to participate in *in vivo* imaging assessment with carotid MRI prior to surgery. Team members performing eligibility screening are bound to uphold data protection requirements. A physician trained in Good Clinical Practice oversees screening activity and ultimately determines eligibility.

### Plaque biobanking

Plaque material is collected directly from the operational room during endarterectomy immediately after plaque excision. Details on sample handling and processing are provided in the **Supplementary Methods**. Each sample is dissected into 5 mm-long segments centered around the most-diseased segment (MDS) exhibiting the highest degree of atherosclerotic burden, as represented by plaque thickness (**Suppl. Fig. I**). MDSs undergo *ex vivo* MRI scanning (**Supplementary Methods, Suppl. Table I**)^33^ and histological processing for morphological plaque characterization, while the surrounding sections are flash-frozen and transferred to long-term storage (−80°C) for omics analyses. The central segments are then decalcified and embedded in paraffin for histological processing. Stainings of serial tissue sections are performed with H&E, Picrosirius Red (for collagen), and immunohistochemistry using anti-alpha-smooth-muscle-actin (αSMA; smooth muscle cells) and anti-CD68 antibodies (macrophages). Plaque features are annotated and quantified from whole-slide scans following a standard protocol (**Supplementary Methods**). The extracted features include total plaque area, lipid core area, calcification area, neovascularization, intraplaque hemorrhage, fibrous cap thickness, macrophage and smooth muscle cell (SMC) content.

### Blood biobanking

Peripheral blood is collected from study participants prior to surgery. Serum and plasma aliquots are kept in long-term storage (−80°C) for further analyses, while whole blood is used for DNA extraction. The detailed protocols are available in the **Supplementary Methods**

### Demographic and clinical data collection

Study participants undergo a structured interview (**Suppl. Table II**) followed by inspection of their electronic health record files. We record detailed information on sociodemographic data, cardiovascular risk factors, biosampling, indication for surgery, medical history, current medications, and the most recent laboratory assessments from clinical routine (**Suppl. Table III**).

### In vivo imaging

We collect vascular and brain imaging studies from the electronic records of the study participants. Specifically, we export pseudonymized imaging stacks in DICOM format from the most recent pre-surgery carotid CTA and brain MRI. The scans are acquired pre-surgery during routine clinical assessment and scanner brand and parameter settings vary across patients.

Eligible participants are additionally invited to a pre-surgery high-resolution MRI of the carotid arteries and brain in the context of an imaging study nested within AtherOMICS (**Supplementary Methods**). The imaging protocol is based on a previous study^34^ and includes time-of-flight MR angiography (TOF-MRA), pre- and post-contrast black-blood T1-, PD- and T2-weighted sequences with fat suppression. Scans are performed on a 3T device (Magnetom Prisma, Siemens Healthineers, Erlangen, Germany) using a routine 32-channel head coil and two 4-channel, special-purpose carotid coils. A physician assesses whether contraindications for MRI are present and obtains separate written, informed consent from the participant. Participants unwilling or unable to undergo MRI can still participate in the biobank through collection of plaque material, blood samples, and clinical data.

### Omics profiling

Fresh frozen plaque samples, patientserum and plasma samples are stored for further omics analyses. To date we have pursued single-nuclei RNA sequencing (snRNAseq) and affinity-based proteomics and plan to expand to genotyping, bulk RNA sequencing, and spatial transcriptomics (**Supplementary Methods**). Plaque tissue snRNAseq, was conducted through Singleron’s SCOPE-Chip™ platform. Proteomic profiling was performed using Olink’s Explore 3072™proximity extension assay. Briefly, stabilized lysates from frozen plaque sections were incubated with DNA barcodes tagging specific proteins from predetermined panels prior to PCR barcode amplification.

## Results

### Patient cohort

As of November 16, 2025, 246 patients have been recruited to the AtherOMICS biobank (**Fig. 2**). Recruitment started in August 2022 with patients undergoing carotid endarterectomy and was expanded in March 2024 to include patients undergoing femoral endarterectomy (**Suppl. Fig. IIA**). Of the participants, 212 underwent carotid endarterectomy for either asymptomatic (38.2%) or symptomatic (61.8%) carotid stenosis, while 34 patients underwent femoral endarterectomy for advanced peripheral artery disease. Among patients with symptomatic carotid stenosis, the most common qualifying event was ischemic stroke (48.9%), followed by transient ischemic attack (24.4%), amaurosis fugax (12.2%), and central retinal artery occlusion (8.3%) (**Suppl. Fig. IIB**).

**Figure 2.**
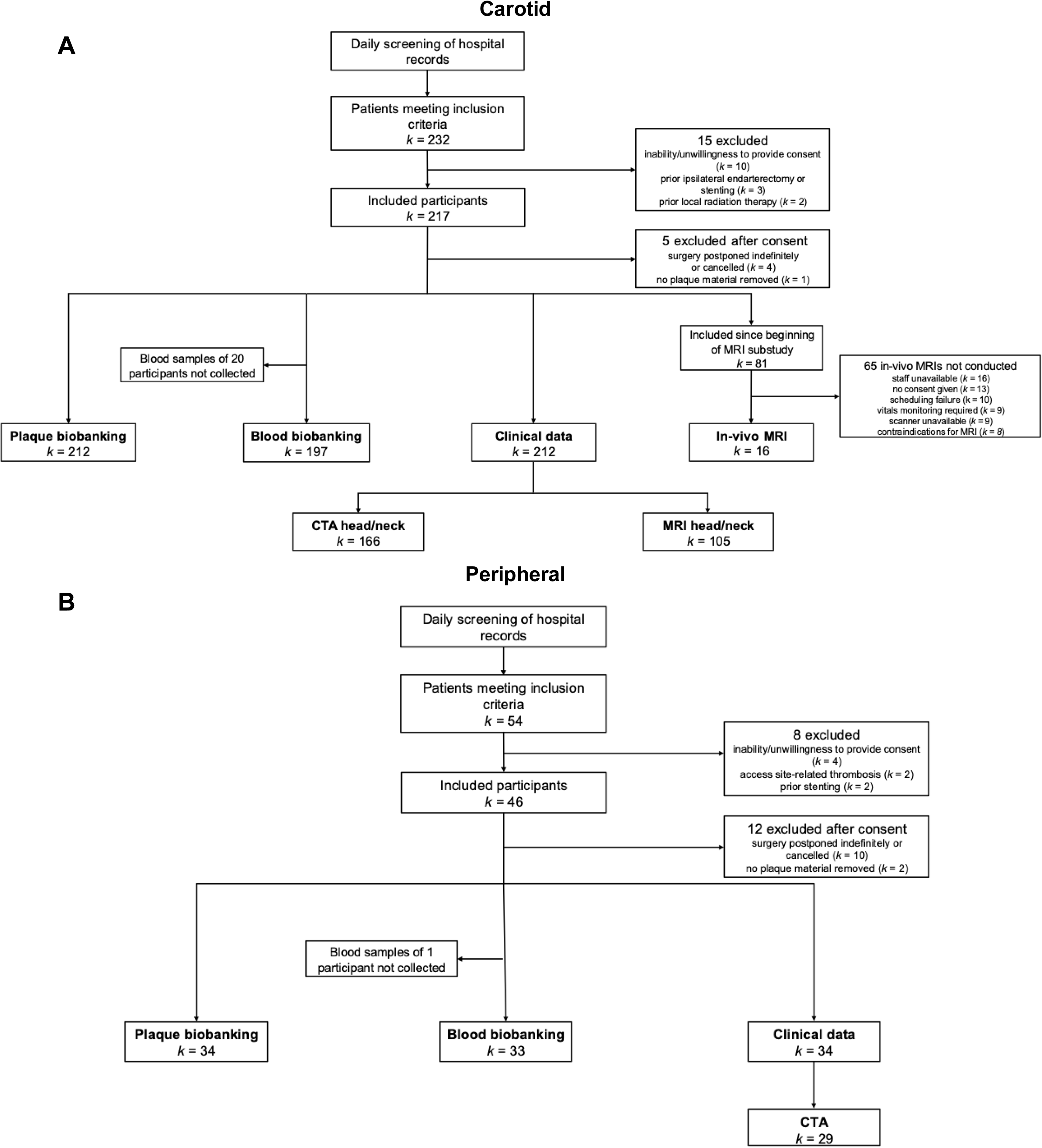
Study flowchart. Shown is the inclusion process for patients undergoing endarterectomy surgery of either the carotid (A) or femoral (B) artery.

Baseline characteristics of carotid endarterectomy patients are summarized in **Table 1**. As expected, there was a high prevalence of vascular risk factors with 77%, 69%, and 25% reporting a history of hypertension, hypercholesterolemia, or diabetes, respectively. Furthermore, 35%, 26%, and 16% of the patients undergoing carotid endarterectomy had a history of coronary artery disease, prior stroke, or peripheral artery disease, respectively.

**Table 1.**
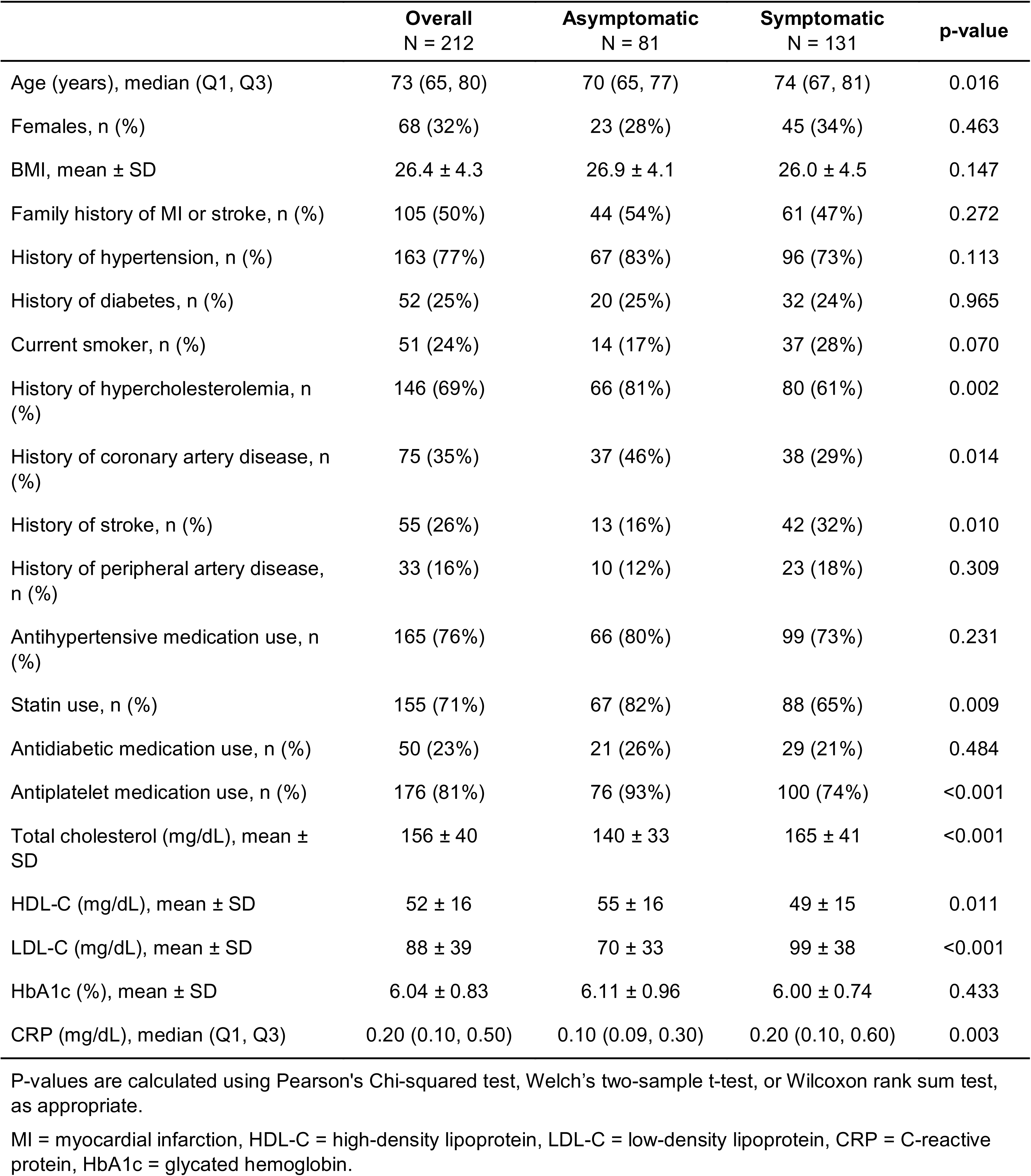
Baseline characteristics of recruited carotid endarterectomy patients in the AtherOMICS study.

Baseline characteristics of the recruited femoral endarterectomy patients are presented in Suppl. Table IV.

### Histopathological plaque characterization

Collected plaques undergo initial processing until flash-freezing within a median of 40 [IQR 33 to 50] minutes after removal (**Suppl. Fig. IIC**). All excised plaques undergo *ex vivo* MRI and histological assessment for morphological characterization (**Fig. 3A**). Macrophage and SMC contents are quantified via automated segmentation of CD68 and αSMA-positive areas in immunohistochemical stainings (**Fig. 3B-C**). Morphological features, including lipid core size, intraplaque hemorrhage, calcification, neovessel density, and fibrous cap thickness, are manually segmented and annotated across histological and *ex vivo* MRI images (**Fig. 3D-I**). Across all assessed features, intraplaque hemorrhage area was significantly larger in plaques from symptomatic patients (asymptomatic 5.05 ± 11.69% vs. symptomatic 10.02 ± 10.75%, n = 33 vs. 54, p = 0.0012; **Fig. 3H**).

**Figure 3.**
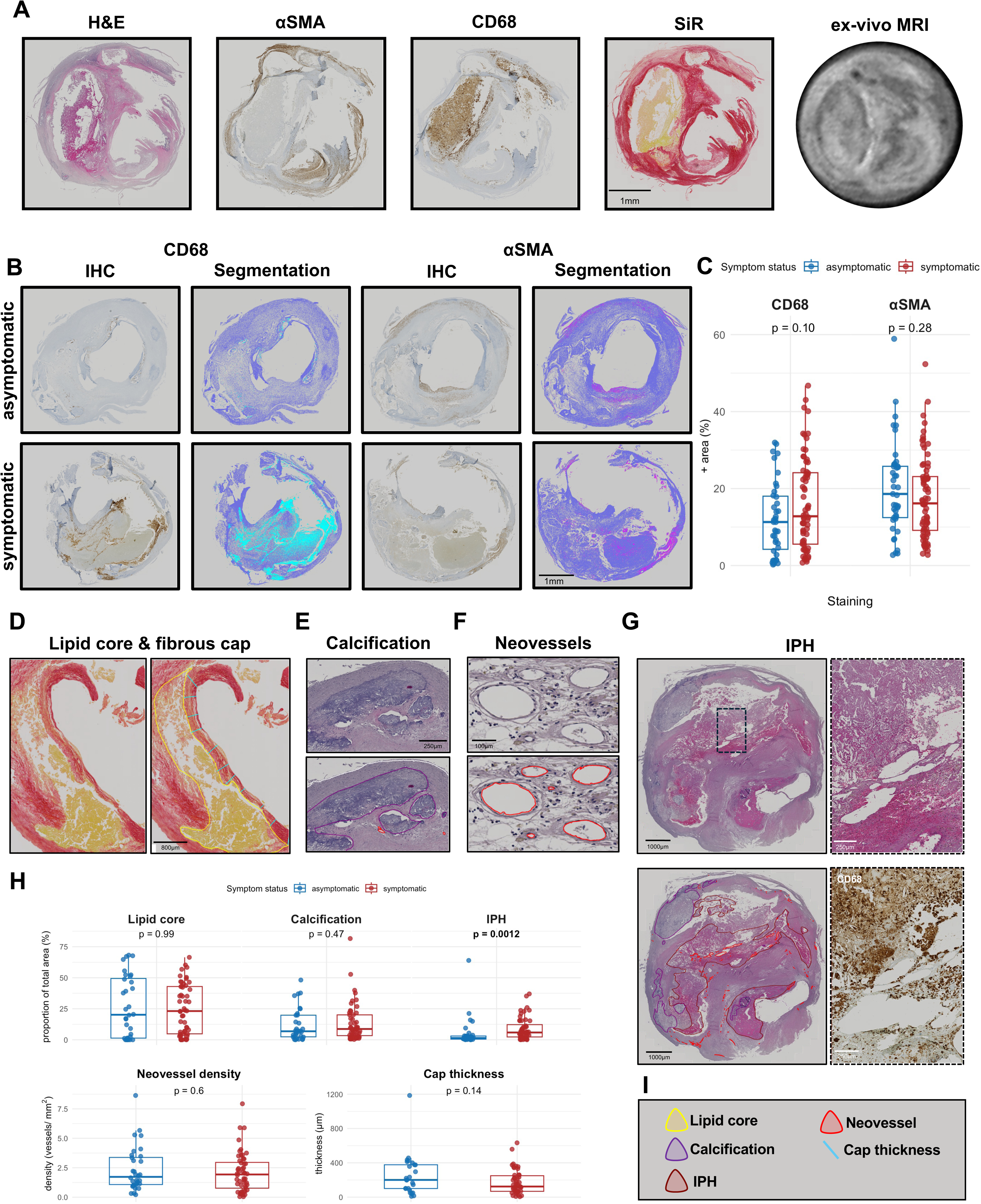
Histological assessment of carotid atherosclerosis in AtherOMICS. **A.** Plaque cross-section across multiple ex-vivo imaging modalities. From left to right: Hematoxylin & eosin (H&E) for general histology, anti-α-smooth-muscle actin (αSMA) for smooth muscle cells, anti-CD68 for macrophages, Sirius Red (SiR) for collagen content and lipid cores, T2*-weighted ex-vivo MRI for morphology. Scale bar represents 1mm. **B.** Quantification of immunohistochemical stainings through automated feature segmentation. Original whole-slide image (IHC) is displayed next to the result of the segmentation algorithm (“Segmentation” panels). In CD68 segmentation, light blue indicates positive signal. In αSMA, purple indicates positive signal. Dark blue indicates negative signal in both stainings. Scale bar represents 1mm. **C.** Box-and-whiskers-plot of CD68 & αSMA-positive areas relative to total plaque area in asymptomatic (CD68: n = 39; αSMA: n = 41) and symptomatic (CD68: n = 68; αSMA: n = 72) participants. Blue color represents asymptomatic, red color symptomatic participants. Data points indicate individual values and the upper and lower boundaries of the boxes indicate Q3 and Q1, respectively. The median is represented through a bold line. Whiskers extend to 1.5 x IQR above/below their respective quartiles. p-values are provided above. Mann-Whitney U (CD68) and unpaired, two-sided Student’s t-test (αSMA). **D-G.** Representative images depicting plaque vulnerability features. Bottom panels (D: right panel) display examples of manual plaque feature annotation in whole-slide scans prior to quantification. Scale bars represent 800µm (D), 250µm (E), 100µm (F), 1000µm (G, left-hand panels) and 250µm (G, right-hand insets). **H.** Box-and-whiskers plots comparing plaque vulnerability features in plaques from asymptomatic (n = 33) and symptomatic (n = 54) participants. p-values considered statistically significant at α < 0.05 are provided in bold. Mann-Whitney U test. **I.** Graphical legend for the annotation line colors used in **D-G**.

### Proteomics and transcriptomics

Plaque segments and peripheral blood samples (serum and plasma) are stored for omics analyses. To date we have performed snRNA-seq on 17 carotid plaque samples, as well as proximity extension assay proteomics (Olink Explore 3072) for 88 paired carotid plaque and plasma samples. snRNAseq revealed a total of 19,091 nuclei across the 17 samples, of which 35% were identified as nuclei of macrophages, 33% SMCs, 13% endothelial cells, 11% T cells, and 7% neutrophils (**Fig. 4A**). Our snRNAseq data points towards macrophages as the predominant immune cell type. While in line with findings from a recent study,^35^ prior data from single-cell RNAseq analyses had suggested T cells as the predominant leukocyte subset.^36^ These diverging results might be related to differences in the viability of immune cell subsets across different protocols.^37^

**Figure 4.**
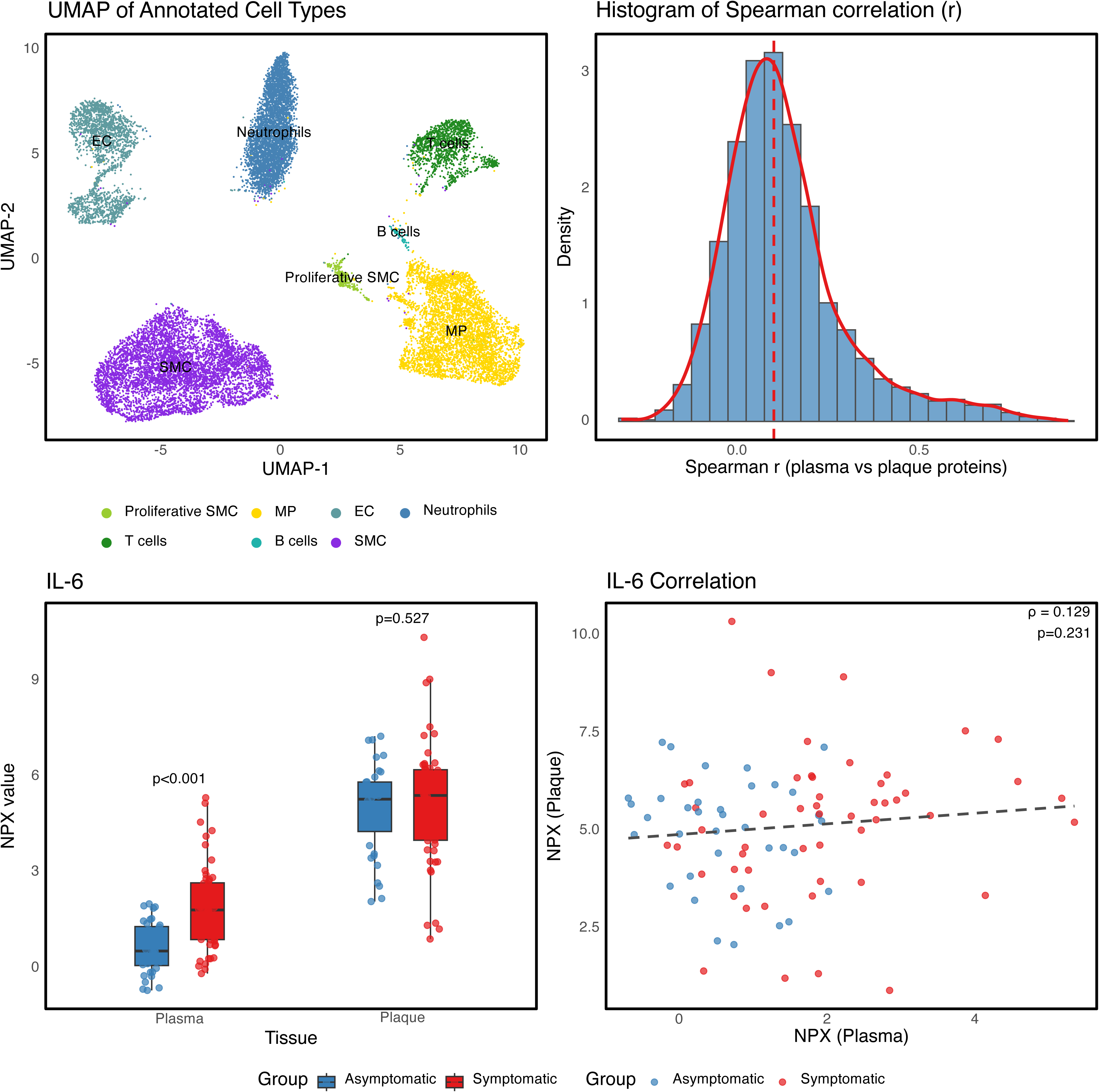
Single-nuclei transcriptomic and proteomic profiling of plaque and plasma samples from carotid endarterectomy patients. **A.** UMAP plot of plaque tissue single-nuclei RNA sequencing data from 17 participants (8 asymptomatic, 9 symptomatic). MP = macrophages, EC = endothelial cells, SMC = Smooth muscle cells. **B.** Histogram and density curve of Spearman correlation coefficients (ρ) between paired plasma and plaque samples computed across 2,810 proteins for 88 carotid endarterectomy patients, overlaid with a density curve. The dashed red line indicates the median correlation coefficient (ρ = 0.10). **C**. Box plots for plaque and plasma interleukin-6 (IL6) normalized protein expression (NPX) values in asymptomatic (blue) and symptomatic (red) patients. P-values are provided above the boxes. Student’s t-test. **D.** Scatterplot of the plasma and plaque NPX values across the 88 sample pairs.

Proteomic analysis of 88 matched plaque and plasma samples revealed the presence of 2,841 distinct proteins. The median Spearman correlation between plaque and serum protein levels for these 2,841 proteins was ρ = 0.10 (IQR 0.02 – 0.20), although 214 proteins showed a correlation of ρ > 0.4 (**Fig. 4B**). Plasma levels of IL-6, a commonly used surrogate of vascular inflammation, were significantly upregulated in patients with symptomatic compared to asymptomatic plaques (p < 0.001; **Fig. 4C**). There was no significant correlation between plaque and plasma levels (Spearman’s ρ = 0.13, p = 0.23, **Fig. 4D**).

### Imaging

We have collected pre-surgery head and neck CTA stacks from 102 patients and performed 9 carotid and brain MRI scans as part of the imaging sub-study nested within AtherOMICS (**Fig. 5**), which began imaging participants in August 2024. We noted that plaque features observed in CTA imaging, such as calcification, were also detectable in *ex vivo* imaging techniques, such as MRI and histology (**Fig. 5A-B**). Similarly, *in vivo* MRI augmented the characterization of non-calcified, soft plaque tissue in CTA (**Fig. 5C-D**). *In vivo* MRI of four symptomatic and 12 asymptomatic patients detected intraplaque hemorrhage in 9 out of 16 carotid arteries (**Fig. 5D**). The quantification of previously described, CTA-derived plaque features revealed no significant differences in total plaque thickness and soft plaque thickness at the level of the index lesion, remodeling index, calcification location, and total calcification volume between asymptomatic and symptomatic patients. There was a statistically significant difference in mean calcification density (asymptomatic 780 ± 322 HU vs. symptomatic 1009 ± 405 HU, n = 32 vs. 71, p = 0.002) (**Fig. 5E**).

**Figure 5.**
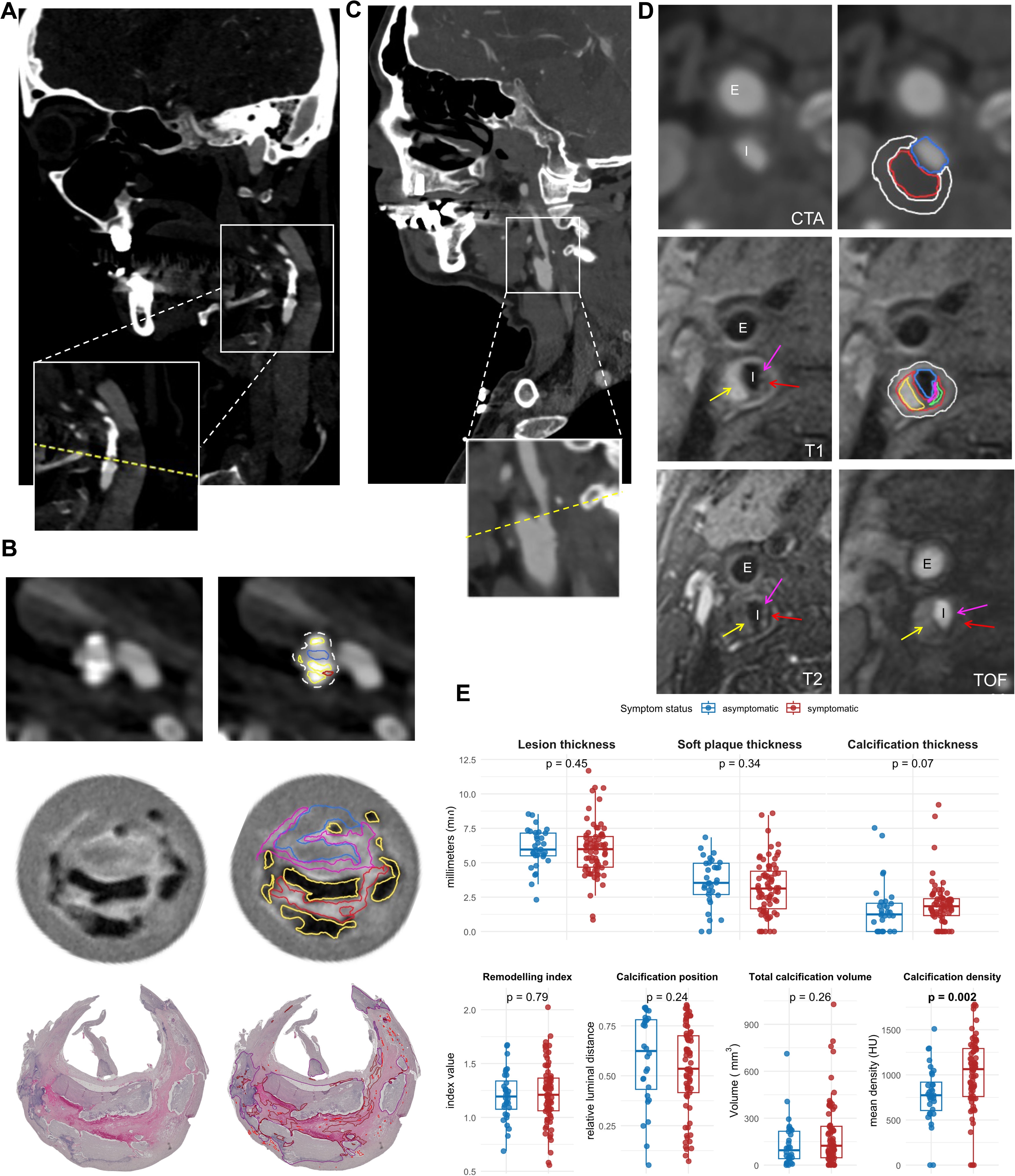
Multi-modal *in vivo* and *ex vivo* carotid plaque imaging in AtherOMICS. **A**. Computed tomography angiography (CTA) in sagittal plane showing the right carotid artery of a female patient presenting with sudden-onset left upper limb motor deficit prior to inclusion into the AtherOMICS study. The yellow dashed line within the inset indicates the location of the axial plane in subpanel B. **B.** Presentation of calcficiation across multiple imaging modalities. Left column displays the original image, right column annotated structures. From top to bottom: Axial-plane CTA, T2-weighted ex-vivo MRI, H&E histology. Annotations: White dashed line denotes outer vessel wall, blue line the lumen, yellow (H&E: purple) line calcification, purple line (H&E: not represented) fibrous cap, red line soft plaque (H&E: intraplaque hemorrhage). **C.** CTA in sagittal plane showing the right carotid artery of a male patient presenting with sudden-onset left upper limb sensorimotor deficit prior to inclusion. The yellow dashed line within the inset indicates the location of the axial plane in subpanel D. **D.** Side-by-side comparison of axial CTA and in-vivo MRI imaging. “E” indicates the external, “I” the internal carotid artery. The outer vessel wall is delineated in white, lumen in blue, soft plaque in red, intraplaque hemorrhage and fibrous cap (MRI sequences) in yellow and purple, respectively. In T1, T2 and time-of-flight angiography (TOF), the purple arrow points to the fibrous cap, the green arrow to a lipid core, and the yellow arrow to intraplaque hemorrhage on the dorsolateral side of the artery. **E.** Box-and-whiskers plots comparing CTA-derived plaque features in asymptomatic (n = 28 - 32) and symptomatic (n = 69 - 71) participants. Student’s t-test (Soft plaque thickness, remodeling index) and Mann Whitney U test (all others).

## Discussion

Here, we present the study protocol and pilot data from AtherOMICS, a biobanking study of patients undergoing endarterectomy surgery that bridges deep plaque phenotyping with blood biobanking, *in vivo* non-invasive imaging and clinical data collection. By integrating these data types, we aim to identify key mechanisms driving plaque progression, understand molecular phenotypes associated with destabilization, and propose non-invasive biomarkers for the detection of vulnerable plaques.

AtherOMICS is well-positioned to address these goals. To elucidate the mechanisms underlying plaque biology, we first aim to map molecular quantitative trait loci (molQTLs) active within atherosclerotic lesions and link them to genetic loci associated with atherosclerotic cardiovascular outcomes. While hundreds of genomic signals have been detected for cardiovascular disease outcomes in genome-wide association studies,^38, 39^ underlying mechanisms remain unknown.^40^ Lead variants located in non-coding genomic regions are assumed to influence gene expression, however, functional genomic studies studying tissue-specific expression are lacking.^41^ To this end, we will genotype samples from the AtherOMICS cohort to identify variants associated with molecular plaque features. Integrating these findings with existing genomic resources for atherosclerotic traits and endpoints^42^ will enable the discovery of specific, genetic drivers of plaque progression and destabilization. Second, we aim to characterize molecular signatures associated with plaque vulnerability. Histopathology has been essential for characterizing atheroprogression, identifying morphological stages of lesion development and substructures linked to plaque instability.^43, 44^ However, molecular features related to unstable plaques remain poorly defined. By linking clinical data and histology with plaque omics profiles, AtherOMICS provides a unique opportunity to investigate these associations. Third, to discover novel biomarkers of symptomatic atherosclerotic disease, we will examine how plaque histological and molecular features relate to *in vivo* imaging phenotypes and to omics profiles in peripheral blood. By aligning imaging and histology data, we aim to characterize how vulnerable plaque substructures appear in noninvasive diagnostics (**Fig. 5**). In parallel, correlating blood-based omics profiles with plaque features will enable the identification of circulating biomarkers predictive of clinical events such as stroke or myocardial infarction.^45^ Together, these efforts can support the development of noninvasive tools for assessing plaque stability and forecasting adverse cardiovascular outcomes.

AtherOMICS has specific limitations. First, endarterectomy patients typically exhibit advanced stages of atherosclerotic disease, which is reflected in a low number of less advanced plaques in the study and thus limits the interpretability of findings for less advanced atherosclerosis stages. Second, it does not follow up on patients after endarterectomy surgery, meaning that recurrent cardiovascular and cerebrovascular events are not captured. Third, despite ongoing recruitment, several omics analyses, typically requiring very large sample sizes, are still underpowered. Fourth, compared to the biobanking and clinical data components of the study, fewer patients agree to participate in the *in vivo* MRI substudy, thus limiting the pace of inclusion. Fifth, the monocentric study design results in an overrepresentation of urban-dwelling individuals of European ancestry.

In conclusion, AtherOMICS integrates deep multi-omic profiling of human atherosclerotic plaques with conventional histopathology, clinical phenotyping, blood biobanking, and *in vivo* plaque imaging. This biobanking effort has the potential to contribute novel insights into mechanisms underlying human atheroprogression, reveal the molecular signatures of plaque vulnerability, and develop non-invasive biomarkers of plaque instability.

## Conflict of interest

ST received consulting fees from Quanterix unrelated to this work. MKG received consulting fees from Tourmaline Bio, Inc., Pheiron GmbH, and Gerson Lehrman Group, Inc. unrelated to this work.

## Financial Support

The AtherOMICS study is funded by the German Research Foundation’s Emmy Noether Programme (GZ: GE3461/2-1, ID 512461526 to M.K.G.) the Munich Cluster for Systems Neurology (SyNergy; EXC 2145, ID 390857198 to M.K.G.), the Fritz Thyssen Foundation (Ref. 10.22.2.024 MN to M.K.G.), the Hertie Foundation (Hertie Network of Excellence in Clinical Neuroscience, ID P1230035 to M.K.G.), and the Vascular Dementia Research Foundation.

## Supporting information

STROBE checklist

## Data Availability

All data produced in the present study are available upon reasonable request to the corresponding author.

## Acknowledgements

We thank Fanni Magdane Boldoczki (Institute for Stroke and Dementia Research, ISD) for her assistance in developing the *ex vivo* MRI protocol, Nadja Sachs (Department for Vascular and Endovascular Surgery, TUM Klinikum, Technical University of Munich) for sharing expertise in plaque processing and biobanking, Andrea Stadie and Tanja Beer (both ISD) for conducting *in vivo* MRI scans, as well as Philip Melton and Karin Waegemann (both ISD) for their assistance in the implementation of the blood biobanking protocol.

## Notes

### Competing Interest Statement

The authors have declared no competing interest.

### Author Declarations

The Ethics Committee of the Faculty of Medicine of the Ludwig-Maximilians-Universitüt (Ethikkommission bei der Medizinischen Fakultät der LMU München) in Munich, Germany (protocol numbers: 121-09, 22-0135, 23-0772).

### Summary of Updates

New Fig. 2 (study flow diagrams) added. Inclusion numbers & data in Table 1, Fig.3 Suppl. Table IV, Suppl. Fig II updated. Wording revised for conciseness.

