## Supplemental Material for "Multi-omic profiling of atherosclerosis: protocol and pilot data for the AtherOMICS biobank"

**Supplementary Materials**

Luka Živković^1^, Roya Batool^1^, Julian Louma^1^, Paulo Vinicius Gil Alabarse^1^, Yingle Li^1^, Lanyue Zhang^1^, Stefan Mayrhofer^1^, Anushree Ray^1^, Mohamad Ali Antabi^1^, Panagiotis Zangas^1^, Jana Mattar^1^, Anna Kopczak^1^, Andreas Schindler^2^, Paul Reidler^3^, Yaw Asare^1^, Steffen Tiedt^1,4,5^, Lars Kellert^4^, Barbara Rantner^6^, Martin Dichgans^1,5,7^, Nikolaos Tsilimparis^6^, Marios K. Georgakis^1,4,8^

^1^ Institute for Stroke and Dementia Research (ISD), University Hospital, LMU Munich, Munich, Germany

^2^ Institute of Neuroradiology, University Hospital, LMU Munich, Munich, Germany

^3^ Department of Radiology, University Hospital, LMU Munich, Munich, Germany

^4^ Department of Neurology, University Hospital, LMU Munich, Munich, Germany

^5^ Munich Cluster for Systems Neurology (SyNergy), Munich, Germany

^6^ Department of Vascular Surgery, University Hospital, LMU Munich, Munich, Germany

^7^ German Centre for Neurodegenerative Diseases (DZNE), Munich, Germany

^8^ Program in Medical and Population Genetics and Cardiovascular Disease Initiative, Broad Institute of MIT and Harvard, Cambridge, MA, USA

**Supplementary Methods**

**Initial visit**

Once a patient’s eligibility is confirmed during screening, they are contacted in person by a member of the study team, who explains the nature of the study using the study consent forms. Patients are then informed about their rights as participants in a clinical study. If patients express interest in participating and once all questions have been addressed, they are asked to sign the consent forms. Copies are provided for the participants’ own records. Consent forms signed and dated by both parties are later stored at the research institution under restricted access. Once written informed consent is obtained, the team member conducts a structured interview for cardiovascular risk factors (**Suppl. Table I**).

**Plaque biobanking**

*Initial processing*

Plaque material is collected in a 50mL falcon tube filled with sterile phosphate-buffered saline (PBS) and transported to the processing facility on ice. The time between removal of plaque material from the affected artery to placement on ice is recorded. A randomly generated pseudonym is assigned to the sample for later reference. For processing, the plaque is photographed and then sectioned as follows (**Suppl. Fig. I**): First, a 5mm section containing the most diseased segment (MDS) in the internal carotid artery as well as 5mm marginal sections proximal and distal to the MDS are identified by morphological assessment and documented on the plaque outline. Then, the plaque is cut into the respective sections using scissors or a scalpel. The MDS is transferred to a tube and submerged in 4% paraformaldehyde (PFA) solution to be used for *ex vivo* MRI and histopathological analysis. Twenty-four hours after initiating PFA fixation, the MDS is removed and washed for 1h under lukewarm running water. Thereafter, the plaque is kept in 70% ethanol in an Eppendorf tube to undergo *ex vivo* MRI. Then, the plaque is decalcified at 4°C using a Tris-buffered 200mM EDTA solution at a pH of 8.0. Daily checks are performed where the section is morphologically assessed for successful decalcification, after which the plaque is transferred to temporary storage in a 70% ethanol solution. The remaining plaque segments are flash frozen by submersion in liquid-nitrogen-cooled 2-methyl butane and stored at -80°C for subsequent omics analyses. The time between placement on ice in the operating room and flash-freezing is again recorded. Per protocol, the sum of table-to-ice-time and ice-to-flash time should not exceed 60 minutes to minimize RNA and protein degradation (**Suppl. Fig. II**).

*Histological processing*

After dehydration and embedding in paraffin, MDS segments are sent out for histological processing at the Core Facility Pathology & Tissue Analytics, Helmholtz Munich, Germany. There, serial sections of 5µm thickness are taken from the MDS in 1mm intervals. The sections undergo stainings with H&E, anti-alpha-smooth-muscle-actin (ɑSMA) immunohistochemistry (IHC) for smooth muscle cells, anti-CD68 IHC for macrophages, and Picrosirius Red for collagen. IHC is performed using standard diaminobenzidine staining. Slides are scanned at high resolution with a microscope (Carl Zeiss NTS Ltd., Oberkochen, Germany). The resulting whole-slide images are stored in the .CZI format. For analysis, whole-slide images of up to three sections are automatically split into scenes containing one section each and imported into QuPath (v.0.4.4).^1^ A set of unstained, unimaged slides is stored for future stainings.

*Immunohistochemistry analysis*

For cellular composition measurements (CD68 and αSMA), QuPath’s segmentation feature is used to manually detect the positive region within the image that can be identified as objects in a hierarchical method below the annotations. These objects can be assigned defined colors in QuPath that classify DAB-stained area (immunostaining – positive, “brown”), immunostaining-immunostaining – negative) and background region devoid of tissue, based on the predictions at the region of interest at the same time. QuPath’s random trees classifier feature consists of multi-class classification tasks which assign each pixel of the image to one of the three categories, e.g., “positive”, “negative” and “ignore” (no plaque region) and are used for model training. Thereby, values for immunostaining-positive area and remaining plaque area (both in µm^2^) are obtained. This workflow is implemented throughout the images of immunostained sections and resulting quantification data are collected in spreadsheets for later analysis. αSMA sections are quantified in moderate and CD68 samples in high resolution due to differences in color intensities.

*Plaque feature annotation strategy*

Staff with experience in vascular histology and unaware of sample identity annotate plaque features using the geometry tools provided within the software. Sections are only annotated if they contain at least 50% of plaque circumference and are devoid of out-of-focus areas.

Total plaque area is determined through measuring the area of the entire plaque tissue captured in the H&E cross-section and serves as a normalizing parameter for the other measurements. If large parts of the cross-section are not represented in H&E but other stainings at an adjacent level within 5-10µm, the other staining is used to impute total plaque area.

Neovasularization is determined by annotating each visible neovessel in any given H&E section. Neovessels are distinguished from other, irrelevant structures through judging the shape (mostly round, oval or elongated), observing elongated endothelial cells surrounding the lumen, and the presence of red blood cells inside the lumen.^2^ The shape of the lumen is overlain with a geometric annotation to capture both total number and total area of neovessels per section.

Intraplaque hemorrhage (IPH) is typically detected in H&E and definitively identified using corroborating signatures in Sirius Red and CD68 stainings. IPHs typically present as either fresh, with large quantities of clearly delineable red blood cells present within plaque tissue with strong CD68-postive signal, or old IPH, with sparse or degraded-appearing red blood cells, clear borders between the light-pink-colored hemorrhage area and surrounding plaque tissue, and a homogenous structure that obscures the underlying plaque matrix.^3, 4^ Occasionally, mixed forms can be observed that result from re-hemorrhaging at the site of a previous IPH.^5^

Calcification can be observed in H&E stainings. Despite the plaque tissue undergoing decalcification, the borders of previously fully calcified islets can usually be clearly delineated and inferred using matched *ex vivo* MRI, which is performed before decalcification. Sheet-calcified areas and calcified nodules exhibit a dark purple to deep blue staining in H&E, whereas microcalcifcations display blue granular deposits within the extracellular matrix that is best contrasted from the surrounding, uncalcified tissue at larger magnification levels.^5, 6^

Lipid cores are detected and characterized in Sirius Red stainings. Their main distinguishing features are clefts resulting from dissolved cholesterol crystals, dark yellow (lipid-rich) matrix, the absence of collagen fibers in later lesional stages and outlines of cellular debris throughout the core.^5, 7, 8^ As the unstable core tissue is often impacted during processing, especially in ruptured plaques, parts of the lipid core are at times not accounted for in Sirius Red sections. In these cases, if the plaque geometry is unaffected and the surrounding plaque tissue permits the inference of original shape and size, the lipid core area is reconstructed using directly adjacent sections and *ex vivo* MRI to confirm.

Likewise, fibrous caps are examined in Sirius Red stainings. Caps present as tightly packed collagen fibers featuring a deep red color and separate underlying lipid cores from the lumen.^9, 10^ Fibrous cap thickness is defined as the shortest rectangular distance between the point of luminal contact of a fibrous cap and the underlying lipid core, which is a mandatory criterion for cap thickness measurement.

*Ex-vivo MRI*

MRI scanning is undertaken in a 3T small animal scanner (nanoScan PET/MRI, Mediso, Münster, Germany) using a 30 mm diameter birdcage coil within one to four days after excising specimen. Sequences were based on previously published data with minor changes.^11^ Quantitative T1/T2/T2* mapping sequences (qT1, qT2, qT2*) and multi-contrast fast-spin echo (FSE) sequences including T1, T2, T2*, and proton-density (PD) weighted images are acquired in axial orientation. A multi-inversion recovery (IR) fast spin-echo (FSE) sequence with 12 inversion times (TIs) (30, 50, 80, 100, 300, 400, 500, 700, 800, 1000, 1200, 1300 ms) is used for T1 mapping; a multi-echo spin-echo sequence with 14 echoes ranging from 10 ms to 140 ms with 10 ms steps is used for T2 mapping; and an interleaved multi-echo GRE sequence with 8 echoes ranging from 2.19 ms to 17.59 ms with 2.2 ms steps is used for T2* mapping. Scanning parameters are summarized in **Suppl. Table I**. The proton density values of plaque components are also quantified by normalizing the signal intensities on PD-weighted images, with the signal intensities of the healthy wall set to 1 as a reference.

**Blood biobanking**

Peripheral blood is collected in two 9mL EDTA-coated tubes and one 7.5mL serum tube (S-Monovette™, Sarstedt, Nümbrecht, Germany). The serum and one EDTA tube are maintained in an upright position for 30mins at room temperature, then centrifuged at 2000g, 15°C for 10mins to separate serum and plasma. Aliquots of 8 x 300µL for each serum and plasma are created in barcoded tubes, loaded onto 8x12 racks and scanned into an internal database. The samples are stored at -80°C to be later used for omics analyses. The second EDTA-coated plasma tube is stored at 4°C for DNA extraction.

*DNA extraction*

DNA is extracted from participant plasma using commercially available NucleoSpin Blood XL tube and reagent kits (Macherey-Nagel, Düren, Germany) as well as the supplied protocol. Briefly, 250µL of 20µg/mL Proteinase K suspension are added to 5mL of EDTA plasma and incubated for 15mins at 56°C after adding the supplied buffer and vortexing the solution. When cooled to room temperature, 100% ethanol is added and the entire solution is loaded onto the supplied filter columns. After centrifuging twice at 4000g for 3mins and removing the flow-through at both iterations, the remaining sample is washed twice under centrifugation using the supplied wash buffer. Elution is performed using 500µL of 70°C warm BE buffer and incubated for 3mins, then centrifuged for another 2mins at 4000g. The eluate is then collected and DNA concentration is measured using a NanoDrop 2000 spectrophotometer (Thermo Fisher, Waltham (MA), USA). Samples are then catalogued in an internal DNA database and stored at -80°C for later use.

**Single-nuclei RNA sequencing**

Frozen plaque sections of 150-500mg for 32 plaques were sent to Singleron Biotechnologies (Cologne, Germany) to perform single-nuclei RNA sequencing. Briefly, for library preparation after tissue dissociation and cell lysis nuclei were extracted, counted, then nuclei concentrations are adjusted to 1x10^6^/mL. Nuclei were suspended in a mix with reverse transcriptase reagent, containing template oligoprimers, and reverse transcriptase. The solution was loaded onto a chip (SCOPE-chip®) and surrounded by oil surfactant. Nuclei then form droplets with a bead gel containing unique molecular identifiers (UMIs) using a microfluidic double cross connection isolation system. Formed droplets were collected, cDNA fragments reverse transcribed, and gel beads subsequently dissolved, releasing the UMI-tagged amplification products. Barcoded fragments were pooled for library construction, then sequenced aiming for 6000 nuclei and 35000-50000 reads. Quality control was performed by examining tissue-, nuclei-, cDNA and library yield and fragmentation. Nuclei from a total of 17 plaques passed quality control and were successfully sequenced.

Raw count matrices were processed in R (v4.3.2) with Seurat (v4.3).^12^ Nuclei with <150 or >6,000 detected genes, <150 or >50,000 UMIs, or >20 % mitochondrial reads were removed. Counts were variance-stabilised with SCTransform,^13^ regressing out mitochondrial content. Doublets were identified with DoubletFinder^14^ and excluded. Samples were integrated with Seurat’s SCT workflow (1,500 shared features; 15 principal components). Integrated data were embedded with PCA and UMAP, and graph-based Louvain clustering (resolution 0.1) delineated major cell populations. Clusters were annotated using marker genes for cell types, including SMCs, macrophages, T cells, endothelial cells and neutrophils obtained from published single-cell RNA sequencing data.^15^

**Proteomics**

We have performed proteomic characterization of 88 matched plaque-plasma pairs. To guarantee sample integrity, both blood and plaque samples destined for proteomics analysis were stored at -80°C as described above. RIPA lysis buffer with added complete Mini protease inhibitor cocktail (Sigma-Aldrich, St. Louis (MI), USA) was used for cell lysis. Plaque samples were thawed on ice, homogenized in the lysis buffer, and centrifuged to remove debris. The resulting supernatant was stored at -80°C until further use. Plaque lysates were subjected to SDS-PAGE and subsequent Coomassie Blue staining, whereafter gels were inspected visually for signs of protein degradation. Pseudonymized plaque lysates and plasma aliquots were shipped on dry ice to Helmholtz Munich, Germany, on 96-well PCR plates specified by the assay provider. Proteomic profiling was performed using Olink's Explore 3072 panel, a proximity extension assay platform that utilizes DNA-conjugated antibodies and subsequent DNA amplification and sequencing to determine protein abundance.^16^ The resulting Ct values were converted into normalized protein expression (NPX), a surrogate marker of protein concentration, through adjusting for inter-plate correction factors. Finally, mean NPX values from a total of 2841 proteins in both plaque and plasma were compared between symptomatic and asymptomatic patients. In addition, associations between plaque and plasma NPX values for any given protein were examined through calculating Spearman’s rank correlation coefficients.

**Clinical Data**

Sociodemographic and anthropomorphic data are acquired from hospital records. Cardiovascular risk factors are assessed through a structured interview (**Suppl. Table II**). Carotid endarterectomy patients are considered to be symptomatic if they report sudden-onset focal neurological symptoms consistent with cerebrovascular events ipsilateral to the stenosed carotid artery within the past six months, or if such events are found in the patient records.^17, 18^ A NASCET value obtained during clinical routine is extracted from past radiological or ultrasound examinations. “Cardiovascular medication” is defined as the current medications upon admission to the hospital that fall under one or more of the following categories: (1) Antihypertensive, (2) Antidiabetic, (3) Lipid-modifying agent, (4) Antiplatelet agent or anticoagulant, (5) Diuretic. For clinical laboratory assessments, most recent values from the moment of surgery are extracted. It is not uncommon for the individual assessments (**Suppl. Table III**) to be performed at different timepoints; therefore, the most recent leukocyte count & C-reactive protein (CRP) measurements are extracted if they occurred up to 14 days prior to surgery. Total cholesterol, high-density lipoprotein (HDL) and low-density lipoprotein (LDL) measurements are extracted if they occurred up to 28 days before surgery.

***In vivo* imaging data**

*in vivo MRI substudy*

MRI scans are performed on a 3T device (Magnetom Prisma, Siemens Healthineers, Erlangen, Germany) using a routine 32-channel head coil and two 4-channel, special-purpose carotid coils. The imaging protocol has been previously described.^19, 20^ Briefly, it includes time-of-flight MR angiography (TOF-MRA), black-blood T1-, PD- and T2-weighted sequences with fat suppression. Study pseudonym, sex and age are recorded and imaging stacks archived in a dedicated clinical imaging server inside the research facility.

We aim to pursue the integration of contrast-enhanced T1w sequences. Patients without known intolerance to gadolinium and no evidence of severely impaired renal function (GFR ≤ 30 ml/min/kg) will also be administered gadolinium-DTPA-BMA (Gadobutrol, Bayer-Schering, Leverkusen, Germany) at a dosage of 0.1mL/kg and at a rate of 3mL/s, 5 minutes before post-contrast T1w imaging.

*CTA annotation strategy*

CTA imaging stacks are exported from the clinical documentation system and loaded into 3DSlicer (v. 5.6.2)^21^ for further processing. The slice with the most severe luminal narrowing in the carotid artery is defined as the index lesion level. If there is no substantial variation in the degree of stenosis between adjacent slices, vessel wall morphology is also considered, and the slice with maximum plaque burden as determined by plaque diameter is selected as the index lesion. Total plaque, soft plaque and calcification thickness measurements are taken from the index lesion slice. Calcification location is similarly measured at the index lesion level and expressed as relative luminal distance of the center of the largest calcification islet. This relative distance is determined by dividing the distance of the calcification islet from the lumen and the distance from the lumen to the outer vessel outline.^22^ The remodeling index was calculated as the ratio of the cross-sectional lumen area at the most stenotic site to the lumen area of the proximal reference.^23^ To obtain total calcification volume, calcified areas were manually delineated throughout the entire plaque. Mean calcification density was defined as the average Hounsfield unit value within the region spanning from the lumen to the outer wall contour throughout the entire plaque volume.^24^

| **Sequence** | **T1w** | **T2w** | **T2*w** | **PDw** | **qT1** | **qT2** | **qT2*** |
| --- | --- | --- | --- | --- | --- | --- | --- |
| **Sequence type** | SE 2D | FSE 2D | GRE 2D | FSE 2D | Multi IR FSE | Multi Echo SE 2D | Interleaved Multi Echo GRE 2D |
| **NEX** | 6 | 8 | 4 | 4 | 1 | 1 | 4 |
| **TR [ms]** | 701 | 12980 | 25 | 3883 | 24711 | 3856 | 578,52 |
| **TE [ms]** | 8,39 | 49,74 | 1,6 | 8,29 | 6,77 | 10 - 140 (10 ms steps) | 2.19 -  17.59 (2.2 steps) |
| **Flip angle** | 90 | 90 | 30 | 90 | 90 | 90 | 90 |
| **Th/gap [mm]** | 0.4/0.00 | 0.4/0.00 | 0.4/0.00 | 0.4/0.00 | 1/0.20 | 1/0.20 | 1/0.20 |
| **FOV [mm]** | 30x30 | 30x30 | 30x30 | 30x30 | 30x30 | 30x30 | 30x30 |
| **Matrix** | 256x256 | 256x256 | 256x256 | 256x256 | 150x150 | 150x150 | 256x256 |
| **Resolution [um]** | 117x117 | 117x117 | 117x117 | 117x117 | 200x200 | 200x200 | 117x117 |
| **Sp** |  |  |  |  | 0.20/0.20 | 0.20/0.20 | 0.12/0.12 |
| **Number of slices** | 60 | 60 | 60 | 60 | 18 | 18 | 18 |
| **Scan time** | 0:35:57 | 0:17:44 | 0:32:26 | 0:11:00 | 1:29:47 | 0:09:42 | 0:19:56 |
| **Bandwidth [Hz]** | 50080 | 50080 | 390625 | 50080 | 50080 | 50080 | 150240 |
| **ETL** |  | 24 |  | 6 | 8 |  |  |
| **TI [ms]** |  |  |  |  | [30, 50, 80, 100, 300, 400, 500, 700, 800, 1000, 1200, 1300] |  |  |

**Supplementary Table I. Scanning parameters for *ex vivo* MR imaging of plaque tissue.**

NEX: No. of averages; TR: Repetition time; TE: Echo time; Th/gap: slice thickness per gap between slices; FOV: field of view; Matrix: No. of columns x No. of rows; ETL: Echo train length; TI: Inversion time; SP: Spatial resolution.

**Supplementary Table II. Structure and contents of the cardiovascular risk factor portion of the interview at initial visit.**

| **No.** | **Item** | **Wording** | **Recorded answers** |
| --- | --- | --- | --- |
| 1 | Self-reported family history of myocardial infarction or stroke | “Do you know if any of your parents, grandparents or siblings have suffered a stroke, a myocardial infarction and/or peripheral artery disease?” | Yes/No/Unknown |
| 2 | Self-reported history of hypertension | “Do you have or are you being treated for elevated blood pressure?” |  |
| 3 | Self-reported history of diabetes | “Do you suffer from or are you being treated for diabetes?” |  |
| 4 | Self-reported history of dyslipidemia | “Do you have or are you being treated for elevated cholesterol levels?” |  |
| 5 | Self-reported history of coronary artery disease | “Do you suffer from or are you being treated for coronary heart disease, or have you ever suffered a myocardial infarction?” |  |
| 6 | Self-reported history of stroke | “Have you ever suffered a stroke?” |  |
| 7 | Self-reported history of peripheral artery disease | “Have you ever been diagnosed with peripheral artery disease?” |  |
| 8 | Smoker status | “Do you smoke cigarettes?”  *If yes:*  “How many packs a day do you smoke and at what age did you start?”  *If former smoker:*  “How many packs a day have you smoked before quitting, how many years ago did you quit, and at what age did you start?” | Yes/No |
| 9 | Self-reported height/weight | “How tall are you and how much do you weigh?” | Height in cm, weight in kg |
| 10 | BMI | N/A | Calculated from 9 |

| **General information** | **ASCVD risk survey** | **Clinical information** | | **CV medication *(at presentation)*** | | **Surgery information** | **Biosampling information** | | **Clinical lab values** |
| --- | --- | --- | --- | --- | --- | --- | --- | --- | --- |
| Date of recruitment  Date of birth  Sex  Patient ID  Case ID | See **Suppl. Table II** | Site of surgery | Carotid  Other | Type | Antihypertensive  Statin  Antidiabetic  OAC/APA  Diuretic  Other | Plaque ID  Date of surgery  Table-to-ice time  Ice-to-flash time  No. of sections for histology/omics | Obtained blood sample | Serum  Plasma  Full blood  (DNA) | Total cholesterol  HDL  LDL  HbA1c  CRP  Total blood leukocytes  Blood neutrophils  Blood lymphocytes  Blood monocytes  Blood eosinophils  Blood basophils |
|  |  | Symptomatic stenosis? | Yes/No | Name | e.g. “Atorvastatin” |  | Date & time of blood sample | |  |
|  |  | *if SCAS:* Most recent clinical manifestation | Stroke  TIA  AF RAO  Other  Unknown | Dosage | e.g. “40 mg” |  | Date & time of last food/beverage consumption | |  |
|  |  | NIHSS at presentation *(if available)* | 0-42 | Timing | morning  noon  evening  night  unknown |  | Alcohol intake up to 24 hours prior | Yes  No  Unknown |  |
|  |  | Symptom onset  *(if available)* | Date & time | TWD | Calculated from dosage + timing * 7 |  | Smoking up to 3 hours prior | Yes  No  Unknown |  |
|  |  | Side of surgery | L/R |  | |  | Cold symptoms up to 4 weeks prior | Yes  No  Unknown |  |
|  |  | NASCET value *(if available)* | 50-99% |  |  |  | Body temperature > 38.0°C up to 4 weeks prior | Yes  No  Unknown |  |

**Supplementary Table III. Overview of recorded clinical data.**

SCAS: Symptomatic carotid artery stenosis, ACAS: Asymptomatic carotid artery stenosis, TIA: Transitory ischemic attack, AF: Amaurosis fugax, RAO: Retinal artery occlusion, NIHSS: National Institutes of Health Stroke Scale, NASCET: North American Symptomatic Carotid Endarterectomy Trial, OAC: oral anticoagulant, APA: antiplatelet agent, TWD: total weekly dose, HDL: high-density lipoprotein, LDL: low-density lipoprotein, HbA1c: Hemoglobin A1c, CRP: C-reactive protein.

**Supplementary Table IV**. **Baseline characteristics of femoral endarterectomy patients in the AtherOMICS study.**

|  | **N = 34** |
| --- | --- |
| Age (years), median (Q1, Q3) | 73 (64, 77) |
| Females, n (%) | 11 (32%) |
| BMI, mean ± SD | 24.5 ± 4.9 |
| Family history of MI or stroke, n (%) | 18 (53%) |
| History of hypertension, n (%) | 28 (82%) |
| History of diabetes, n (%) | 7 (21%) |
| Current smoker, n (%) | 14 (41%) |
| History of hypercholesterolemia, n (%) | 24 (71%) |
| History of coronary artery disease, n (%) | 13 (38%) |
| History of stroke, n (%) | 4 (12%) |
| Total cholesterol (mg/dL), mean ± SD | 150 ± 33 |
| HDL-C (mg/dL), mean ± SD | 60 ± 20 |
| LDL-C (mg/dL), mean ± SD | 76 ± 33 |
| HbA1c (%), mean ± SD | 6.01 ± 0.69 |
| CRP (mg/dL), median (Q1, Q3) | 0.2 (0.09, 1.00) |

MI = myocardial infarction, HDL-C = high-density lipoprotein, LDL-C = low-density lipoprotein, CRP = C-reactive protein, HbA1c = glycated hemoglobin.

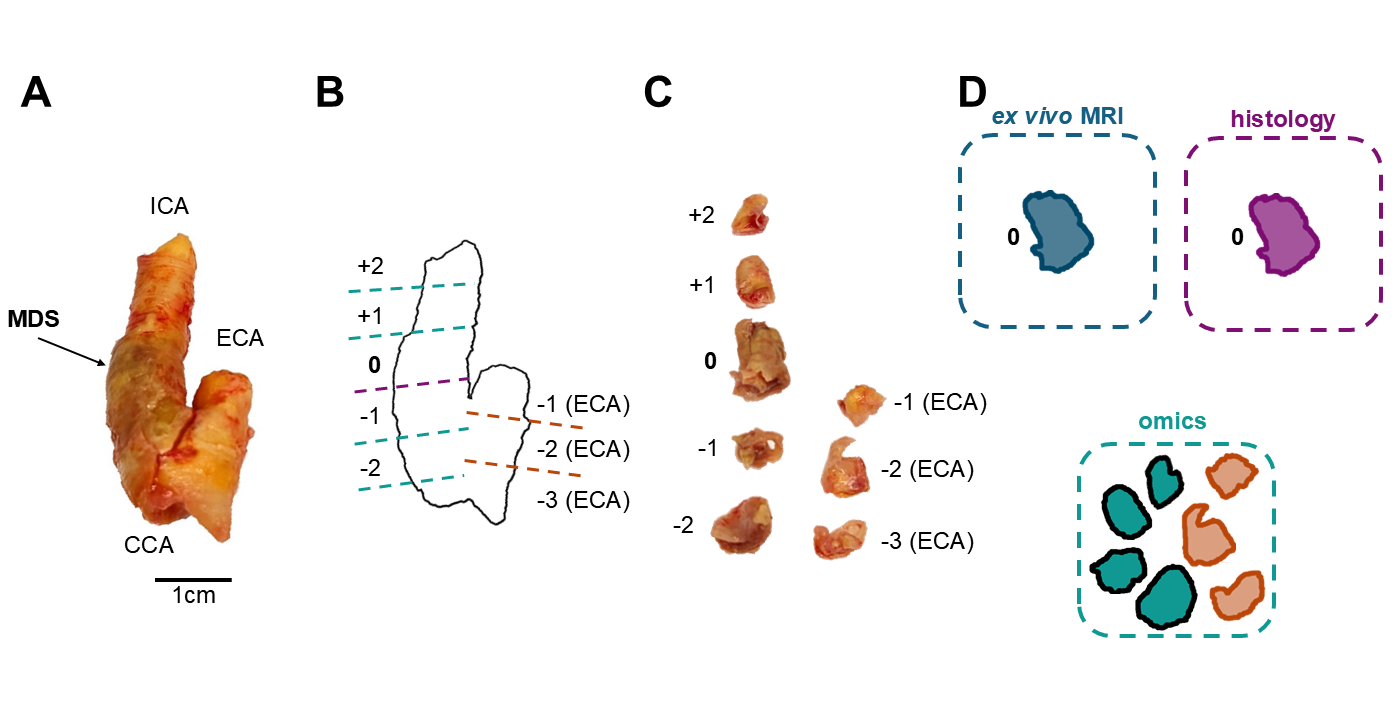

**Supplementary Figure I. Preparation of plaque segments**. (A) Exemplary plaque from the biobank, photographed prior to cutting. CCA: Common carotid artery, ICA: Internal carotid artery, MDS: Most diseased segment, ECA: External carotid artery. (B) The plaque outline is used to plan incisions on paper. The MDS is labelled as “0”; other segments are labelled as “-2”, “-1”, “+1”, “+2” in the direction of blood flow. ECA-derived plaque material is labelled separately. (C) The resulting sections are arranged and photographed once again. (D) The MDS (“0”) is submerged in PFA for 24 hours as described above, then subjected to *ex vivo* MRI, then processed further for histology. The remaining segments are flash-frozen and stored at -80°C for omics analyses.

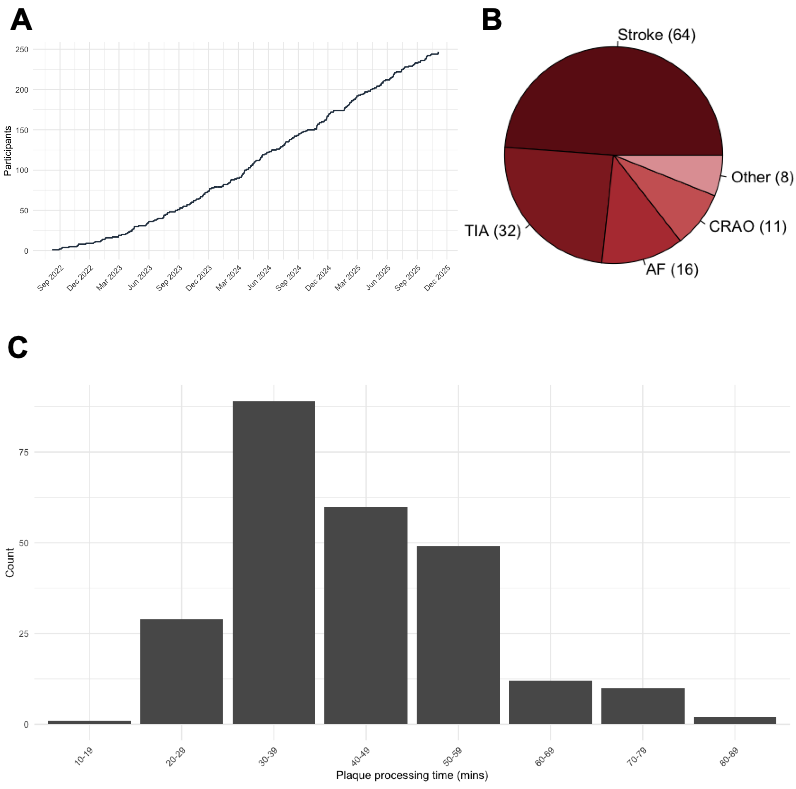

**Supplementary Figure II. A.** Number of included participants over the course of the duration of the study. **B.** Pie chart depicting proportions of cerebrovascular manifestations in symptomatic carotid stenosis patients. The absolute number of cases for each presentation is given in brackets. TIA: Transitory ischemic attack, AF: Amaurosis fugax, CRAO: Central retinal artery occlusion. **C.** Histogram of total plaque processing times, defined as the time between retrieval from the patient in the operating room until flash freezing of plaque sections in liquid nitrogen.
